# Pragmatic, open-label, single-center, randomized, phase II clinical trial to evaluate the efficacy and safety of methylprednisolone pulses and tacrolimus in patients with severe pneumonia secondary to COVID-19: the TACROVID trial protocol

**DOI:** 10.1101/2021.01.09.21249263

**Authors:** X Solanich, A Antolí, N Padullés, M Fanlo-Maresma, A Iriarte, F Mitjavila, O Capdevila, M Molina, J Sabater, J Bas, A Mensa-Vilaró, J Niubó, N Calvo, S Bolivar, R Rigo-Bonnin, L Arregui, C Tebé, P Hereu, S Videla, X Corbella

## Abstract

**Introduction:** Some COVID-19 patients evolve to severe lung injury and systemic hyperinflammatory syndrome triggered by both the coronavirus infection and the subsequent host-immune response. Accordingly, the use of immunomodulatory agents has been suggested but still remains controversial. Our working hypothesis is that methylprednisolone pulses and tacrolimus may be an effective and safety drug combination for treating severe COVID-19 patients.

**Methods and analysis:** TACROVID is a randomized, open-label, single-center, phase II trial to evaluate the efficacy and safety of methylprednisolone pulses and tacrolimus plus standard of care (SoC) versus SoC alone, in patients at advanced stage of COVID-19 disease with lung injury and systemic hyperinflammatory response. Patients are randomly assigned (1:1) to one of two arms (42 patients in each group). The primary aim is to assess the time to clinical stability after initiating randomization. Clinical stability is defined as body temperature ≤ 37.5°C, and PaO2/FiO2 > 400 and/or SatO2/FiO2 > 300, and respiratory rate ≤24 rpm; for 48 consecutive hours.

**Discussion:** Methylprednisolone and tacrolimus might be beneficial to treat those COVID-19 patients progressing into severe pulmonary failure and systemic hyperinflammatory syndrome. The rationale for its use is the fast effect of methylprednisolone pulses and the ability of tacrolimus to inhibit both the CoV-2 replication and the secondary cytokine storm. Interestingly, both drugs are low-cost and can be manufactured on a large scale; thus, if effective and safe, a large number of patients could be treated in developed and developing countries.

**Trial registration number:** NCT04341038 / EudraCT: 2020-001445-39

## 1. BACKGROUND

In December 2019, an emerging disease (COVID-19), caused by a newly identified human coronavirus (SARS-CoV-2), was first recognized in Wuhan, China and spread worldwide [1,2]. The WHO declared the COVID-19 epidemic to be a pandemic on March 12, 2020 [3].

Unfortunately, there are still no proven effective and safe therapies for treating the COVID- 19 illness other than supportive care. Despite the lack of evidence, the urgency of care leads to a large number of severe patients receiving off-label and compassionate use therapies, based on their in vitro antiviral or immunomodulatory properties. Among suggested treatments, the repositioning of older drugs may be a plausible strategy given that their proven safety profile [4,5]. Furthermore, there are several ongoing randomized controlled trials (RCTs) assessing different drug regimens for treating patients with COVID-19 [6].

Of great concern, some COVID-19 patients evolve to fatal lung injury and multi-organ failure due to the serious inflammatory process triggered by the viral infection [7-12]. While the use of some immunosuppressive drugs has been reported to be potentially harmful, other such drugs, paradoxically, have been justified in treating the excessive inflammation triggered by the viral infection [13]. Unfortunately, studies performed following pragmatic randomized controlled trials (pRCT) are still very limited to date and current international recommendations have not taken a position either in favor of or against the use of immunomodulatory therapy in such patients.

Our working hypothesis is that methylprednisolone pulses and tacrolimus might be an effective and safe drug combination strategy for severe COVID-19 patients. Accordingly, given the rapid spread of COVID-19 and the current health emergency worldwide, we performed a proof-of-concept study in a randomized, open-label, single-center, clinical trial to evaluate the efficacy and safety of methylprednisolone and tacrolimus plus standard of care (SoC), versus SoC alone, in severe COVID-19 patients with lung injury and systemic hyperinflammatory syndrome.

The rationale for the present pRCT is based on the fact that glucocorticoids such as methylprednisolone are a mainstay in the treatment of several immune-mediated disorders, with multiple mechanisms of action involving both the humoral and cell-mediated arms of immunity. As for tacrolimus, the rationale for its use is based on its specific mechanism of action leading to an impaired lymphocyte function and a decrease in pro-inflammatory cytokines (PIC) [14]. Interestingly, severe COVID-19 disease presents a fairly similar clinical and cytokine profile to other diseases such as secondary hemophagocytic lymphohistiocytosis (sHLH) [15] or clinically amyopathic dermatomyositis (CADM) associated with anti-melanoma differentiation-associated gene 5 (MDA-5), where calcineurin inhibitors such as tacrolimus or cyclosporine play a central role in its treatment [16]. Furthermore, some human coronavirus (CoV) replication depends on active immunophilin pathways, which can be inhibited by tacrolimus, at low, non-cytotoxic concentrations in cell culture [17]. In this regard, anecdotal case series suggest that tacrolimus may exert a protective effect in solid organ transplanted patients affected by MERS-CoV [13]. Thus, tacrolimus has an immunosuppressive effect but would also block viral replication and may have beneficial impacts on severe COVID-19 lung injury.

## 2. METHODS

### 2.1 Study design

TACROVID is a pragmatical, randomized (1:1), open-label, single-center, phase II clinical trial to evaluate the efficacy and safety of methylprednisolone pulses and tacrolimus plus SoC, versus SoC alone, in severe COVID-19 patients with lung injury and systemic hyperinflammatory syndrome.

### 2.2 Population

Patients are being prospectively recruited and included in the study for subsequent randomization, if they meet all the inclusion criteria and there is no reason for exclusion.

#### Inclusion criteria

1. COVID-19 infection confirmed by nasopharyngeal RT-PCR,
2. New pulmonary infiltrates (either by chest X-ray or computerized axial tomography),
3. Respiratory failure (PaO2 / FiO2 <300 or satO2 / FiO2 <220),
4. High analytical inflammatory parameters (CRP >100 mg/L, and/or D-Dimer >1000 µg/L and/or Ferritin >1000 ug/L).

#### Exclusion criteria

1. Critically ill patients with life expectancy ≤ 24h,
2. Glomerular filtration ≤ 30 mL/min/1.73m^2^,
3. Leukopenia ≤ 4000 cells/µL or other conditions that cause immunosuppression,
4. Concomitant potentially serious infections,
5. Contraindication for the use of corticosteroids or tacrolimus according to the Summary of Product Characteristics,
6. Known hypersensitivity to any of the study drugs, their metabolites, or formulation excipient,
7. Previous participation in a RCT in the last 3 months.

All patients (or a legal representative) have to provide informed consent (ICF) prior to initiation of the study procedures.

### 2.3 Setting

The TACROVID trial is being conducted at Bellvitge University Hospital (BUH), a 750-bed tertiary care public hospital in Barcelona (Catalonia, Spain). BUH is the reference hospital for high complexity patients from the Southern Area of Catalonia, a region of approximately 2 million inhabitants.

### 2.4 Randomization

On day 0, patients are centrally and randomly assigned using the RedCap computer platform that allows data collection and patient randomization. Patients are not stratified by baseline variables. Participants are randomized in a 1:1 ratio into one of two study groups: methylprednisolone pulses, tacrolimus and SoC, or SoC alone.

### 2.5 Treatment

Patients are being randomly assigned to one of the following arms:

1. <u>Experimental:</u> methylprednisolone pulses of 120 mg/day are administered on 3 consecutive days (if not previously administered) and immediate release tacrolimus is administered twice daily until clinical stability is achieved. Tacrolimus dosing is individualized through therapeutic drug monitoring (TDM) to achieve blood trough levels of 8-10 ng/mL. In addition, patients in the experimental arm can receive standard of care (SoC) for their management in accordance with treating physicians.
2. <u>Control (SoC)</u>: SoC includes measures of supplemental oxygen and respiratory support, fluid therapy, antipyretic treatment, postural measures, low molecular weight heparins, and may also include other treatments, whether antiviral or immunosuppressive drugs, or those necessary at the discretion of the treating physician, except for cyclosporine and/or tacrolimus.

The experimental drugs start immediately after having been randomly assigned to this group. The experimental treatment is discontinued after “*clinical stability*”, which is achieved after fulfilling all of the following criteria for 48 consecutive hours: body temperature ≤ 37.5°C; PaO2/FiO2 > 400 and/or SatO2/FiO2 > 300; and respiratory rate ≤ 24 rpm.

Experimental treatment is also discontinued if the included patient has a severe or potentially severe infection, required invasive mechanical ventilation, extracorporeal membrane oxygenation (ECMO), or has any serious medication-related adverse event (of special interest refractory HTA, decrease of more than 50% of the GFR compared to the baseline, or ventricular tachycardia).

### 2.6 Study visits

All patients are followed from day 0 through day +56 or death. The planned visits are: 1) randomization visit (day 0) in which the patient is screened and eventually included in the study; 2) Hospitalization visits that are variable according to the period of admission; 3) Visits on day +28 ±3 and on day +56 ±3 from randomization. These visits are face-to-face to evaluate whether patients present a relapse or worsening of the disease, as well as the presence of adverse events or mortality. Specific procedures that are performed on each visit are detailed in Table 1 and the evaluation methods section.

**Table 1.** Flow-chart of the trial showing the procedures that will be conducted in each visit

|  | Randomization visit (day 0) | Hospitalization visits (days n) | Visit 28 ± 3 days after the start of treatment | Visit 56 ± 3 days after the start of treatment |
| --- | --- | --- | --- | --- |
| Inclusion / exclusion | x |  |  |  |
| Informed consent | x |  |  |  |
| Randomization | x |  |  |  |
| Demographics<br>Comorbidities | x |  |  |  |
| COVID-19 clinical data | x |  |  |  |
| Vital signs | x | x | x | x |
| 8-point Ordinal scale | x | x | x | x |
| Analysis (Hematology, Chemistry, Coagulation) | x | x | x | x |
| Tacrolimus levels |  | x |  |  |
| Cytokines | x | x | x | x |
| Viral load (blood) | x | x | x | x |
| Nasopharyngeal SARS-CoV-2 PCR | x |  | x | x |
| Clinical stability | x | x | x | x |
| Lung tests (x-ray, CT scan, chest US, 6MWT, functional tests) | x | x (1) | x (1) | x |
| Study treatment | x | x | x | x |
| Concomitant medication | x | x | x | x |
| AE registration | x | x | x | x |

(1) according to the attending physician

### 2.7 Evaluation Methods

#### 2.7.1 Clinical parameters

The randomization visit (day 0) includes the collection of demographic variables [date of birth, sex, ethnicity], weight and height, clinical data [symptoms onset date, hospital admission date, clinical manifestations, radiological characteristics of pulmonary infiltrates], comorbidities [smoking and alcohol consumption, Charlson index], functional and cognitive assessment [Barthel and Pfeiffer index respectively], 8-point Ordinal scale (https://clinicaltrials.gov/ct2/show/NCT04280705), vital signs [temperature, blood pressure, heart rate, respiratory rate, oxygen assessment and supply (type of oxygen mask and FiO2, non-invasive or invasive ventilator support devices, or ECMO, PaO2/FiO2 and/or SatO2/FiO2), and conscious level using the Glasgow Coma Scale], as well as concomitant medication. With all these data we automatically calculate body mass index (BMI), the Pneumonia Severity Index (PSI) and CURB-65 index, and determine the extent of organ failure (SOFA score). The hospitalization visits include the collection of vital signs, 8-point Ordinal scale, adverse events and mortality. Visits on day +28 ±3 and +56 ±3 include vital signs, ordinal scale, relapse or worsening of the disease, adverse events and mortality.

#### 2.7.2 Tests and other procedures

A blood analysis is performed before starting the study treatment with the following biomarkers: 1) hematology (hemoglobin, hematocrit, platelet count, absolute lymphocyte count, leukocyte count with differential) assessed using Sysmex^®^ XN2000 analyzer (Sysmex Europe GmbH; Norderstedt; Germany); 2) basic chemistry (creatinine, albumin, ALT, total bilirubin) and inflammatory parameters (ferritin, D-dimer, CRP, LDH) measured with Cobas^®^ 6000/8000 analyzer (Roche Diagnostics^®^, Risch-Rotkreuz, Switzerland), which has spectrophotometry and immunochemistry modules with electrochemiluminescent detection; and 3) coagulation (activated partial thromboplastin time, prothrombin time and fibrinogen) using ACLTOP^®^ 550 analyzer (Werfen^®^; Barcelona, Spain). Furthermore, all patient samples are collected, processed and stored at randomization, and weekly after randomization by the BUH-IDIBELL Biobank following standard operating procedures. During hospitalization, a blood analysis is performed 3 times a week with the same parameters as used at randomization. Tacrolimus levels are measured using a method based on ultra-high-performance liquid chromatography coupled with tandem mass spectrometry (HPLC-MS/MS) [18].

The QTc interval is evaluated using an ECG before randomization, and an electrocardiogram 3 times a week is recommended during hospitalizations. All pulmonary tests performed during hospitalization at the discretion of the treating physician are being collected (chest x-Ray CT scan). A chest CT scan, the six-minute walk test (6MWT) and functional respiratory tests on day +56 ±3 follow-up visit are performed.

In addition, a follow-up oropharyngeal SARS-CoV-2 RT-PCR is performed on day +28 ±3 and +56 ±3 visits. Blood viral load will be evaluated using quantitative RT-PCR to assess the impact of immunosuppressive treatment on viral dynamics. Microbiological tests are being carried out by the Microbiology Department at the BUH.

All samples will be sent to the Immunology Laboratory of the Biomedical Diagnostic Center at the Clinic Hospital in Barcelona to perform a multiple cytokines quantification technique by Luminex^®^. Concentration of the following cytokines will be analysed: IL-6, IL-18, CXCL10, CXCL9, TNF-alpha, IFN-alpha, IFN-beta, IFN-gamma, IL1RA, IL1-beta, IL10 and IL-2R / CD25.

### 2.8 Outcomes

#### 2.8.1 Primary end point

Average of days until clinical stability from treatment randomization

Treatment failure is defined as: 1) patient who does not meet the criteria for clinical stability 56 days after starting treatment; 2) patient presenting with a grade 3 or 4 adverse event attributed to trial treatment; or 3) patient who dies after being included in the clinical trial.

#### 2.8.2 Secondary endpoints

- Days until normalization of each of the clinical parameters (body temperature, PaO2/FiO2 and/or SatO2/FiO2, respiratory rate) from day +1 through day +56.
- Days until normalization of each of the analytic parameters (D-dimer, CRP, ferritin, LDH, IL-6) from day +1 through day +56.
- Number of patients requiring non-invasive and invasive ventilatory support devices during hospitalization.
- Clinical status according to the 8-point Ordinal scale from day +1 through day +56.
- Number of patients who reach a clinical status ≤ 2 after 10 days or hospital discharge, whichever is first.
- Number of patients who reach clinica lstability after 10 days or hospital discharge, whichever is first.
- Value of each of the analytical values (D-dimer, CRP, ferritin, LDH, IL-6) after 10 days or hospital discharge, whichever is first.
- Days with trial experimental treatment.
- Days until hospital discharge.
- Changes in blood quantitative viral load by RT-PCR before start treatment and weekly during hospitalization.
- Changes in expanded cytokine profile before starting treatment and weekly during hospitalization.
- Long-term efficacy (28 and 56 days after trial treatment initiation) measuring whether clinical stability are maintained and the incidence of relapse of COVID-19 illness.
- Description of the radiological abnormalities (chest x-ray, chest CT scan, functional tests) at day 56.
- Adverse events according to their severity and relationship to trial experimental treatment.
- COVID-19 related mortality at day +28 and +56 after randomization.
- All-cause mortality at day +28 and +56 after randomization.

#### 2.8.3 Criteria for withdrawing a patient from the study

Patients must be withdrawn from the study in any of the following situations: 1) any adverse event for which clinicians consider it necessary to withdraw trial experimental therapy; 2) the patient requests to be withdrawn from the study (at any time during the patient’s participation); 3) the principal investigator (PI) considers that there has been a serious protocol violation; 4) lost to follow-up; and 5) pregnancy during the study.

When a patient withdraws from the study, the investigator records the reason/s in the clinical chart and the electronic case report form (eCRF). If the reason for withdrawal is a serious adverse event, the patient must be followed-up until resolution or stabilisation of the event. Patients who withdraw will not be replaced.

### 2.9 Statistical analysis

#### 2.9.1 Determination of sample size

The median time to clinical stability in the control group is expected to be 16 days. If the hazard ratio of clinical stability of control patients in relation to the patients in the experimental group were 0.52, it would be necessary to include 42 patients in each group in order to be able to reject the null equality hypothesis with a power of 80%. The probability of Type I error associated with this hypothesis test is 0.05 and includes 5% of withdrawal.

#### 2.9.2 Statistical considerations

A descriptive analysis of the study variables will be carried out. Continuous variables will be described as mean and Standard Deviation (SD) or as median and range; and categorical variables as absolute frequencies and percentages. The main analysis will be performed when all patients reach clinical stability or failure. If patients are in hospital for 56 days, they will be considered clinical failures. Comparison between average time to achieve clinical stability between the two study groups will be measured using the log-rank test. To quantify the degree of association, hazard ratio will be estimated with a Cox proportional hazards model using a 95% confidence interval.

Efficacy analyses will be performed for the intention to-treat (ITT) population. If a proportion of subjects >10% is detected to have relevant deviations from the protocol (for example, non-compliance with the selection criteria, non-compliance or inability to receive the trial treatment), the per protocol (PP) analysis will exclude those subjects with such deviations. If this group is defined, all exploratory efficacy analyses will be repeated as a sensitivity test.

As additional analysis, a Cox proportional hazards model, adjusted for clinically relevant confounding factors such as age, sex, comorbidities using the Charlson index, indexes to assess the severity of pneumonia (PSI and CURB-65), index of organic dysfunction (SOFA) and inflammatory parameters will be performed. In addition, prespecified subgroup analyses involving sensitivity analysis will be used to evaluate outcomes in patients not receiving invasive mechanical ventilation.

For evaluation of secondary variables, we will calculate unadjusted and adjusted estimations of effect size and the corresponding 95% confidence intervals using linear, logistic or proportional hazards Cox regression. Study adverse events will be described according to their severity and relationship with other treatments, and will be compared between treatment groups. Midway through recruitment, i.e. 21 patients per arm, an intermediate efficacy and safety analysis will be performed. For this, a correction of the type I error will be applied following Lan-DeMets (O’Brien – Fleming) in evaluation of efficacy.

The IDIBELL Biostatistical Unit will perform analysis and analysts will be blind to the treatment received by the patients (intervention vs. usual care). R version 3.6.2 or higher for Windows (R Foundation for Statistical Computing, http://www.r-project.org) will be used for data treatment and analysis.

### 2.10 Monitoring

An electronic case report form (eCRF) has been created using the RedCap computer platform. IDIBELL Clinical Research and Clinical Trials Unit (UICEC-IDIBELL) is carrying out the monitoring of the trial. Regular monitoring is performed by the UICEC IDIBELL according to the ICH GCP. Compliance with the approved protocol and verification of data is evaluated according to applicable regulatory requirements. The key data in the Monitoring Plan are those related to informed consent, primary outcome, mortality and safety data.

### 2.11 Adverse events reporting and quantification

#### 2.11.1 Definitions

⍰ *Adverse event (AE)*: any injury related to medical management (including all aspects of care) that occurs during the patient’s participation in the clinical trial will be considered an adverse event. An adverse event may be related to the study medication or be non- related.
⍰ *Adverse drug reaction (AR): any ‘adverse drug event’ that occurs when the medication is used as directed and in the usual dosage will be considered an adverse drug reaction*.
⍰ Serious AE (SAE) or serious AR (SAR) are considered to be those that at any dose:
  - provoke death (Note: death is a possible evolution of SARS-CoV-2 infection);
  - put patient’s life at risk;
  - require the patient’s hospitalization or the extension of an existing hospitalization.
  - cause permanent or significant disability or incapacity;
  - cause a congenital anomaly or malformation;
  - are considered medically relevant.
- *Adverse drug event of particular interest for the study*: The investigator shall record in the eCRF and communicate to the promoter, the AE that are considered of special interest as soon as possible and no later than 15 days after he becomes aware of them. The AEs that are considered of special interest are:
  - refractory hypertension (defined as poor blood pressure control despite 3 antihypertensive drugs including a diuretic);
  - renal impairment (decrease of more than 50% of the GFR compared to the baseline);
  - ventricular tachycardia.

#### 2.11.2 Reporting

The UICEC-IDIBELL is carrying out pharmacovigilance of the trial. All SAEs (including death), regardless of their relationship with the investigational medications, have to be notified by the investigator within 24h. The investigator has to make the notification via the SAE Notification Form, sending it by email to the pharmacovigilance unit that will review it and, if appropriate, will request additional information from the investigator. The investigator will provide information to the promoter or pharmacovigilance unit whenever requested and, in any case, when its initial evaluation changes regarding severity or causation. Likewise, all additional information regarding the AE, until the end of the study or until its definitive outcome, must be communicated without delay, via follow-up reports following the notification procedure previously described.

In case of a suspected unexpected serious adverse reaction (SUSAR), UICEC-IDIBELL will report to the competent authorities. The sponsor will also report the development update safety report (DSUR) once a year to the ethics committee and the Spanish Agency for Drugs and Health Products (AEMPS).

### 2.12. Ethical considerations

This trial conforms to the Declaration of Helsinki and Good Clinical Practice guidelines, and personal data protection as required by Spanish law (LOPD 3/2018). The protocol and informed consent form (ICF) were approved by the BUH’s Ethical Committee for Drug Research (EC) and by the Spanish Agency for Drugs and Health Products (AEMPS) in March 2020, in accordance with current legislation, Royal Decree 1090/2015 of December 4 and European Regulation 536/2014 of April 16, regulating clinical trials with drugs. The study received the ethics approval by the Research Ethics Committee of Bellvitge University Hospital (AC010/20).

The patient, his or her closest legal or family representative (in case of incapacity due to the severity of the clinical situation) has to accept the ICF. As patients with COVID-19 can infect researchers via the ICF document, patients can consent orally with a witness, and this is documented in the patient’s medical history. If the legal representative or a close relative is in quarantine due to COVID-19 quarantine, informed consent is provided orally by telephone and documented in the patient’s medical history. The written ICF is obtained when the patient or his/her closest legal or family representative are able to give consent, as they are not quarantined.

Methylprednisolone and tacrolimus are commercially available as generic drugs, so no special permissions are required. Both the promoter and the center are respectively responsible for the treatment of patient data and commit to comply with Regulation (EU) 2016/679 of the European Parliament and of the Council of April 27, 2016 on Data Protection (RGPD), as well as with all other laws and regulations in force and applicable (Organic Law on Protection of Personal Data and Guarantee of Digital Rights 3/2018 of December 05). The data collected for the study are identified by a code, so as not to include any information that can identify the patient (name, surname, initials, address, social security number, etc.). Only the study clinician/collaborators are able to relate such data to the patient and his/her medical history. Therefore, a patient’s identity is not revealed to any other person except the health authorities when required, or in the case of medical emergency. Access to patient information is restricted to the attending physicians, the health authorities AEMPS, the Clinical Research Ethics Committee, and personnel authorised by the sponsor when they need to check the data and procedures used in the study, always maintaining confidentiality in accordance with current legislation. If participants wish to know more, they can contact the Promoter’s Data Protection Officer.

#### 2.12.1 Protocol amendments

The protocol was approved by the EC and the AEMPS on March 31, 2020, in a process of authorization adapted to the pandemic situation and based on a protocol synopsis (March 28, 2020). A substantial amendment to the original protocol was submitted to the EC and the AEMPS in accordance with Spanish legislation, and accepted on April 9, 2020. We decided to reduce tacrolimus plasma levels from 10-15 to 8-10 ng/mL and not to maintain corticosteroids by protocol beyond the three pulses of methylprednisolone in the experimental group. In addition, some exclusion criteria were added, such as glomerular filtration ≤ 30 mL / min / 1.73m2, leukopenia ≤ 4000 cells / µL or other conditions that cause immunosuppression, and concomitant and potentially serious infections. Another additional amendment was accepted by the EC and AEMPS on May 25, 2020, including a complete clinical study protocol with new secondary key outcomes (8-point Ordinal scale and radiological abnormalities) that are being analysed in several COVID-19 clinical trials. These outcomes are being prospectively reported in the trial’s eCRF, but were not reported in the protocol synopsis. We do not remove any of the previous key study outcomes.

### 2.13 Publication plans

The trial is currently actively recruiting patients. Completion of patient recruitment is expected for Q3-Q4 of 2020. The sponsor commits itself to publishing the data within 12 months of completion of the study. Results will be analysed and reported in accordance with CONSORT guidelines.

## 3. DISCUSSION

Clinically, COVID-19 patients can evolve into 3 stages of progressive severity. Viral incubation and early establishment of the disease are the predominant components in the first week from the onset of symptoms. In the second stage, viral replication and transition into moderate acute lung involvement are the most important components. The third and most advanced stage manifests as severe pulmonary injury and systemic multi-organ failure, resulting from cytokine storm and systemic hyperinflammatory response. Older age, comorbid chronic conditions, elevated body mass index, lymphopenia, and elevated transaminases, LDH, D-dimer, ferritin, and soluble IL-2 receptor (sIL-2R) on admission are some of the reported risk factors associated with higher mortality [7-12]. From a pathological viewpoint, two different pathological mechanisms appear to coexist in the COVID-19 illness; the first, triggered by the virus itself, and the second, by the host-immune response. Accordingly, a two-step sequence of antiviral and anti-inflammatory drug administration has been proposed, regarding the natural 3-stage evolution of COVID-19 disease [19].

Despite randomized clinical trials (RCTs) being the only way to find effective and safe treatments, very limited available RCTs for treating COVID-19 have been reported to date [20]. Given the emergency situation, most patients are receiving compassionate, unproven therapies to avoid clinical progression into severe advanced stages where host-immune inflammatory response is the most important component [21].

Several studies demonstrate that cells infected by coronavirus (CoV) produce elevated levels of PICs in order to tackle the invading virus. The overproduced PICs may cause immuno- mediated damage to the lungs and other organs, resulting in severe lung injury and systemic hyperinflammatory syndrome [22]. Once this excessive inflammatory response has been triggered, it may self-perpetuate in some patients despite a decrease in the CoV viral load. So, it may be necessary to add “anti-inflammatory” treatments such as corticosteroids, anti-IL-6 or anti-IL-1 inhibitors, Janus kinase (Jak) inhibitors, polyclonal immunoglobulins, etc. to reduce the secondary deleterious inflammatory response triggered by the virus [6].

In this regard, glucocorticoids appear as a key first-line option, especially given their worldwide availability and cost. However, their use has been controversial in patients with SARS-CoV, MERS-Cov or influenza infections. On the one hand, intravenous steroid use has been associated with delayed elimination of coronaviruses in the blood and lungs of patients with MERS-CoV [23] and SARS-CoV [24], and steroids associated with an increased risk of mortality and adverse events in influenza patients [25-27]. On the other hand, a Cochrane review of glucocorticoids as adjunctive therapy in influenza found evidence of low quality due to confounding by indication [28], and there are some series which suggest improvement of severe COVID-19 lung injury after administration of steroids [8,10].

Furthermore, a small retrospective observational study conducted in China [29] suggests the efficacy of tocilizumab (an IL-6 inhibitor) in the treatment of 21 COVID-19 patients with severe pneumonia and high IL-6. But tocilizumab can cause even deeper immunosuppression than steroids, increasing the risk of sepsis, bacterial pneumonia, gastrointestinal perforation, and hepatotoxicity [20]. Another drawback of tocilizumab is its shortage in some hospitals worldwide and its high cost to national health systems. Interestingly, IL-1 blockade has shown particular promise in cytokine storm syndrome and high-dose regimens have been shown safe even in the context of overt sepsis [30]. Emapalumab (anti-IFNγ) is FDA-approved for HLH and may be effective in COVID-19 inflammatory phase. JAK inhibition appears promising; however, the safety of this medication in severe viral infection remains unknown. Accordingly, some ongoing RCTs are studying glucocorticoids and blockage of IL-1, IL-6, and IFNγ in COVID-19 [6]. However, at the end of April 2020, no trials assessing calcineurin inhibitors, whether tacrolimus or cyclosporine, had been registered either at www.clinicaltrials.gov nor EU Clinical Trials Register, thus prompting us to carry out the present investigation.

Given the scarcity of medical reports supporting the use of immunosuppressive therapy in severe COVID-19, the TACROVID trial was initiated on March 29, 2020. The study is a pragmatical, randomized, open-label, single-center, phase II clinical trial to evaluate the effectiveness and safety of methylprednisolone and tacrolimus in combination regimen plus SoC, versus SoC alone, in COVID-19 with severe lung injury and systemic hyperinflammatory syndrome. We decided to consider methylprednisolone pulses for their rapid onset of action, combined with the ability of tacrolimus to inhibit both the multiple PICs, and the SARS-CoV replication in cell cultures. Moreover, the rationale of our study is aligned with an Ovid MEDLINE review article [13] searching for current evidence for immune-suppressing or stimulating drugs to treat COVID-19, which concludes that low-dose methylprednisolone and tacrolimus may have a beneficial impact in such COVID-19 populations.

Tacrolimus has been administered for decades to prevent allograft rejection in transplanted patients as well as to treat patients with severe autoimmune diseases, and it has a well-known safety profile. In addition, it is a low-cost drug and can be manufactured on a large scale. Thus, if tacrolimus is effective in treating the inflammatory process triggered by COVID-19, a large number of patients could be treated in developed and developing countries. Despite the fact that tacrolimus has been shown to be safe for other diseases, we [researchers] are particularly aware of the possible occurrence of associated AE throughout the present TACROVID trial. In this respect, most patients in the advanced stage of the COVID-19 illness are currently given on immunosuppressive treatment and it is well-known that this facilitates certain opportunistic infections, making it difficult to attribute them to one particular drug. Moreover, the drawback of tacrolimus is that it may interact with some antiviral treatments used in COVID-19 (especially Lopinavir). Finally, tacrolimus is started when patients transition into the most severe clinical stage which, in turn, facilitates the occurrence of AE.

Interestingly, in addition to its immunomodulatory effect, tacrolimus has been also reported to show certain in vitro activity against different human coronaviruses, inhibiting viral replication through immunophilins, although its efficacy and safety have not been assessed in clinical practice. For this reason, an interim analysis will be conducted when half of patients (21 per arm) will be recruited in order to measure the basal and weekly viral load, and the potential AE related to immunosuppression.

Regarding the study design, the present trial was not designed as a double-blind trial. This was considered unrealistic given the emergency situation and the extensive workload that it would involve for the hospital pharmacy service. For the above-mentioned reasons, the TACROVID trial was designed as an open study which allows the use of all drugs that treating physicians prescribe, even without clear evidence to support their use. To minimize the impact of an open study, the primary end-point “time to clinical stability”, is based on objective, quantitative measures for 48 consecutive hours: body temperature, and PaO2/FiO2 and/or SatO2/FiO2, and respiratory rate. Likewise, the statistician performing analysis is blind to the treatment that patient receives (experimental vs control).

Cytokine measurement throughout the trial can reveal new available information on COVID- 19 evolution and potential new treatment targets. IFNγ and IL-1β are very relevant cytokine in MAS, but they are not easily assessed in the peripheral blood. CXCL9, a stable chemokine, is a useful surrogate for IFNγ activity in MAS, as are IL-18 and IL-1RA for IL-1β. Thus, in this trial we will not only analyse how the cytokine profile evolves after treatment with tacrolimus, but we will also be able to detect increases in cytokines that can also be treated by other proven specific inhibitors (e.g. IL-18 with Tadekinig alpha, IFNγ with Emapalumab).

In summary, the TACROVID trial assesses the combined use of tacrolimus and methylprednisolone, in addition to the standard of care in patients with severe COVID-19 illness. The aim is to evaluate both the beneficial effect of tacrolimus on controlling viral replication and, in combination with methylprednisolone pulses, modulating the host-immune inflammatory response triggered by the virus. In addition, potential adverse events related to this immunosuppressive therapy are carefully followed-up throughout the investigation. The results of the present trial might encourage further RCTs to assess the efficacy and safety of new antiviral and anti-inflammatory drugs regimens, in sequential combination, at earlier clinical stages of COVID-19.

## Data Availability

All data referred to the manuscript are available upon request to the corresponding author

## ABBREVIATIONS

CoV: Coronavirus
SARS-CoV: Severe Acute Respiratory Syndrome Coronavirus
MERS-CoV: Middle East Respiratory Syndrome Coronavirus
SARS-CoV-2: Severe Acute Respiratory Syndrome Coronavirus 2
COVID-19: Coronavirus disease 2019
SoC: Standard of Care
PaO2: arterial oxygen partial pressure
SatO2: Oxygen saturation
FiO2: fractional inspired oxygen
PIC: pro-inflammatory cytokines
HLH: hemophagocytic lymphohistiocytosis
CADM: clinically amyopathic dermatomyositis
MDA-5: anti- melanoma differentiation-associated gene 5
RT-PCR: Reverse transcription polymerase chain reaction
LDH: lactate dehydrogenase
CRP: c-reactive protein
ALT: alanine aminotransferase
ICF: informed consent form
ECMO: extracorporeal membrane oxygenation
GFR: Glomerular filtration rate
BMI: body mass index
PSI: Pneumonia Severity Index
SOFA score: Sequential Organ Failure Assessment Score
6MWT: six-minute walk test
eCRF: electronic case report form
ARDS: acute respiratory disease syndrome
JAK: Janus Kinase
IL: interleukin
BUH: Bellvitge University Hospital

## Acknowledgments

We wish particularly to thank the patients for their collaboration. Biobank BUH-ICO-IDIBELL (PT17/0015/0024) and UICEC-IDIBELL have the support of the Spanish Platform for Clinical Research and Clinical Trials, SCReN (Spanish Clinical Research Network), funded by the ISCIII – General Subdirectorate for Evaluation and Promotion of Research

## Funding

The TACROVID trial was supported by a special research grant from the Catalan Ministry of Health, Government of Catalonia, through its Department of Research and Innovation (DGRIS), the BioCat research institute, and the Agency for Health Quality and Assessment of Catalonia (AQuAS), geared towards healthcare research centers in the IRISCAT alliance (Health Research and Innovation Institutes of Catalonia) to prevent and treat the Covid-19 disease.

## Contributors

XS, AA, CT, SB, PH and XC took part in the study design, review of the protocol and manuscript writing. NP, MFM, AI, FM, OC, MM, JS, JB, AMV, JN, NC, SB, RRB, and LA took part in the review of the protocol. All authors read and approved the final manuscript.

## Conflicts of Interest

Authors declare no conflicts of interest.

## Notes

### Competing Interest Statement

The authors have declared no competing interest.

### Clinical Trial

NCT04341038

### Funding Statement

No external funding was received

### Author Declarations

The study received the ethics approval by the Research Ethics Committee of Bellvitge University Hospital (AC010/20).

